# SARS-CoV-2 RNAemia with higher nasopharyngeal viral load is strongly associated with severity and mortality in patients with COVID-19

**DOI:** 10.1101/2020.12.17.20248388

**Authors:** Hitoshi Kawasuji, Yoshitomo Morinaga, Hideki Tani, Yoshihiro Yoshida, Yusuke Taekgoshi, Makito Kaneda, Yushi Murai, Kou Kimoto, Akitoshi Ueno, Yuki Miyajima, Koyomi Kawago, Yasutaka Fukui, Miyuki Kimura, Hiroshi Yamada, Ippei Sakamaki, Yoshihiro Yamamoto

## Abstract

**Objective:** This study aimed to determine the frequency of SARS-CoV-2 RNA in serum and its association with the clinical severity of COVID-19.

**Methods:** An analytical cross-sectional study was performed in a single tertiary care hospital and included consecutive patients with confirmed COVID-19. The prevalence of SARS-CoV-2 RNAemia and the strength of its association with clinical severity variables, including required oxygen supplementation, ICU admission, invasive mechanical ventilation, and in-hospital mortality, were examined.

**Results:** Fifty-six patients were included in the study. The median age was 54.5 years, and individuals with RNAemia were older than those without detectable SARS-CoV-2 RNA in serum (78 vs. 50 years; *P* = .0013). RNAemia was detected in 19.6% of patients (11/56) and in 1.0% (1/25), 50.0% (6/12), and 100.0% (4/4) of moderate, severe, and critically ill cases, respectively. Patients with RNAemia required more frequent oxygen supplementation (90.0% vs. 13.3%; *P* < .0001) and ICU admission (81.8% vs. 6.7%; *P* < .0001) and required invasive mechanical ventilation (27.3% vs. 0.0%; *P* < .0001). Among patients with RNAemia, the median viral loads of NP swabs that were collected around the same time as the serum were significantly higher in critically ill cases (5.4 Log_10_ copies/μL [IQR: 4.2–6.3]) than in moderate–severe cases (2.6 Log_10_ copies/μL [1.1–4.5]; *P* =.030) and were significantly higher in nonsurvivor cases (6.2 Log_10_ copies/μL [IQR: 6.0–6.5]) than in survivor cases (3.9 Log_10_ copies/μL [1.6–4.6]; *P* =.045).

**Conclusions:** This study demonstrated a relatively high proportion of SARS-CoV-2 RNAemia and an association between RNAemia and clinical severity. Moreover, among the patients with RNAemia, the viral loads of NP swabs were correlated with severity and mortality, thus suggesting the potential utility of combining serum testing with NP tests as a prognostic indicator for COVID-19 with a higher quality than each separate test.

## Introduction

The kinetics of the SARS-CoV-2 viral load in respiratory airways and other tissues is important for understanding the pathogenesis, course, and management of patients with coronavirus disease 2019 (COVID-19) (1). COVID-19 is most frequently diagnosed from nasopharyngeal (NP) swab samples. Previous studies found that the viral load in NP swab specimens peaks around the time of symptom onset or a few days thereafter and becomes undetectable approximately two or three weeks after symptom onset (2–4); however, there is evidence of prolonged virus detection in elderly patients (2), and previous studies showed that the viral load in the respiratory specimens of symptomatic patients was similar to that of asymptomatic patients (3), thus implying that the viral load in respiratory specimens may not objectively reflect disease severity.

Preliminary data suggest that SARS-CoV-2 RNA may also be detected in plasma and serum (4). However, information regarding the use of other samples to improve patient management is lacking or inconsistent. Serum SARS-CoV-2 viral RNA (termed RNAemia by the authors in a recent study) was detected in 15% of COVID-19 patients (1, 5), but the clinical meaning of SARS-CoV-2 RNAemia detection has not yet been completely elucidated.

In this study, we conducted a retrospective, monocentric cohort study of consecutive COVID-19 patients to investigate the frequency of SARS-CoV-2 RNA in serum and its association with the clinical characteristics and severity of COVID-19.

## Materials and methods

### Study design

Consecutive patients who were admitted to Toyama University Hospital with confirmed COVID-19 from April 13, 2020, to September 28 2020, were tested for SARS-CoV-2 RNA in serum and were included in this retrospective cohort study. NP samples that were collected approximately during admission were included in the study as paired NP samples if residual tested serum were collected within seven days before or after NP collection. Given that viral culture was not performed to determine the viability of the virus, the presence of viral RNA in serum is referred to as RNAemia instead of viremia.

### Data Collection

A retrospective chart review was performed for all individuals in the study to identify basic demographic data, medical history, clinical presentation, and hospital admission. The outcome data collected included the need for oxygen supplementation, admission to an intensive care unit (ICU), invasive mechanical ventilation, and all-cause mortality. Severity was divided into five categories: asymptomatic, mild (symptomatic patients without pneumonia), moderate (pneumonia patients without required oxygen supplementation), severe (required oxygen supplementation), and critically ill (required invasive mechanical ventilation, shock, and/or multiple organ dysfunction). The sample size was based on a convenience set of all available clinical samples without formal power calculation.

### Quantitative reverse transcriptase polymerase transcription assay (RT-qPCR)

Serum samples and NP swabs were collected from all patients, and RNA was extracted. NP swab specimens were pretreated with 500 µL Sputazyme (Kyokuto Pharmaceutical, Tokyo, Japan). After centrifugation at 20,000 × × g for 30 min at 4 °C, the supernatant was used for RNA extraction. A total of 60 µL RNA solution was obtained from 140 µL of the supernatant or 140 µL of the serum by using the QIAamp ViralRNA Mini Kit (QIAGEN, Hilden, Germany) or Nippongene Isospin RNA Virus (Nippongene, Tokyo, Japan) according to the manufacturer’s instructions. The viral loads of SARS-CoV-2 were quantified on the basis of an N2 gene–specific primer/probe set by RT-qPCR according to the protocol of the National Institute of Infectious Diseases of Japan (6). The quality of quantification was controlled using the AcroMetrix COVID-19 RNA Control (Thermo Fisher Scientific, Fremont, CA). The detection limit was approximately 0.4 copies/µL (2 copies/5 µL). When undetectable, viral load data were hypothetically calculated as −0.5 Log_10_ copies/µL.

### Statistical analysis

Continuous and categorical variables are presented as median (interquartile range [IQR]) and n (%), respectively. We used the Mann–Whitney U test, χ^2^ test, or Fisher’s exact test to compare the differences between the patients with and without RNAemia. Data were analyzed using JMP Pro version 14.2.0 software (SAS Institute Inc., Cary, NC, USA).

### Ethics approval

This study was performed in accordance with the Declaration of Helsinki and was approved by the Ethical Review Board of the University of Toyama (approval No.: R2019167). Written informed consent was obtained from all patients.

## Results

Fifty-six patients were included, with a median age of 54.5 years. A total of 24 patients (42.9%) were male, and 8 patients (14.3%) had a Charlson comorbidity index ≥ 2 (Table 1). Hypertension was the most common comorbidity, followed by dyslipidemia, diabetes, and bronchial asthma. Hypertension was significantly more frequent in patients with and without detectable RNAemia. SARS-CoV-2 RNAemia was detected in 11 (19.6%) patients, 9 of whom were admitted to the ICU. Individuals with detectable RNAemia were significantly older than those without RNAemia (78 vs. 50 years; *P* = .0013).

**Table 1.** Demographic and clinical characteristics of 56 patients with paired nasopharyngeal and serum samples tested for SARS-CoV-2.

| Characteristics | Overall<br>patients, n =<br>56 | With<br>RNAemia,<br>n = 11 | Without<br>RNAemia,<br>n = 45 | *P*-value |
| --- | --- | --- | --- | --- |
| Age, median, y | 54.5 | 78 | 50 | .0013 |
| 0–19, n (%) | 4 (7.1) | 0 (0.0) | 4 (7.14) | .58 |
| 20–64, n (%) | 32 (57.1) | 4 (36.4) | 28 (62.2) | .18 |
| $\geq 65$, n (%) | 20 (35.7) | 7 (63.6) | 13 (28.9) | .042 |
| Sex |  |  |  |  |
| Male, n (%) | 24 (42.9) | 5 (45.5) | 19 (42.2) | 1 |
| Presence of symptoms, n (%) |  |  |  |  |
| Asymptomatic | 6 (10.7) | 0 (0.0) | 6 (13.3) | .33 |
| Symptomatic | 50 (89.3) | 11 (100.0) | 39 (86.7) | .33 |
| Mild | 9 (18.0) | 0 (0.0) | 9 (23.1) | .18 |
| Moderate | 25 (50.0) | 1 (9.1) | 24 (61.5) | .0046 |
| Severe | 12 (24.0) | 6 (54.6) | 6 (15.4) | .014 |
| Critically ill | 4 (8.0) | 4 (36.4) | 0 (0.0) | .0014 |
| Comorbidities, n (%) |  |  |  |  |
| Hypertension | 13 (23.2) | 6 (54.6) | 7 (15.6) | .013 |
| Dyslipidemia | 9 (16.1) | 2 (18.2) | 7 (15.6) | 1 |
| Diabetes | 5 (8.9) | 1 (9.1) | 4 (8.9) | 1 |
| Bronchial asthma | 4 (7.1) | 0 (0.0) | 4 (8.9) | .58 |
| Charlson comorbidity index $\geq 2$ | 8 (14.3) | 2 (18.2) | 6 (13.3) | .65 |
| Treatment, n (%) |  |  |  |  |
| Antiviral therapy | 19 (33.9) | 9 (81.8) | 10 (22.2) | .0004 |
| Favipiravir | 17 (30.4) | 8 (72.7) | 9 (20.0) | .0016 |
| Remdesivir | 4 (7.1) | 4 (36.4) | 0 (0.0) | .0009 |
| Antibiotic therapy | 15 (26.8) | 9 (81.8) | 6 (13.3) | <.0001 |

The numbers of asymptomatic, mild, moderate, severe, and critically ill cases were 6, 9, 25, 12, and 4, respectively. Figure 1 shows the proportion of patients with detectable RNAemia stratified by severity. The proportions of patients with RNAemia in moderate, severe, and critically ill cases were 1.0% (1/25), 50.0% (6/12), and 100.0% (4/4), respectively, but there were no cases in asymptomatic and mild cases.

**Fig 1.**
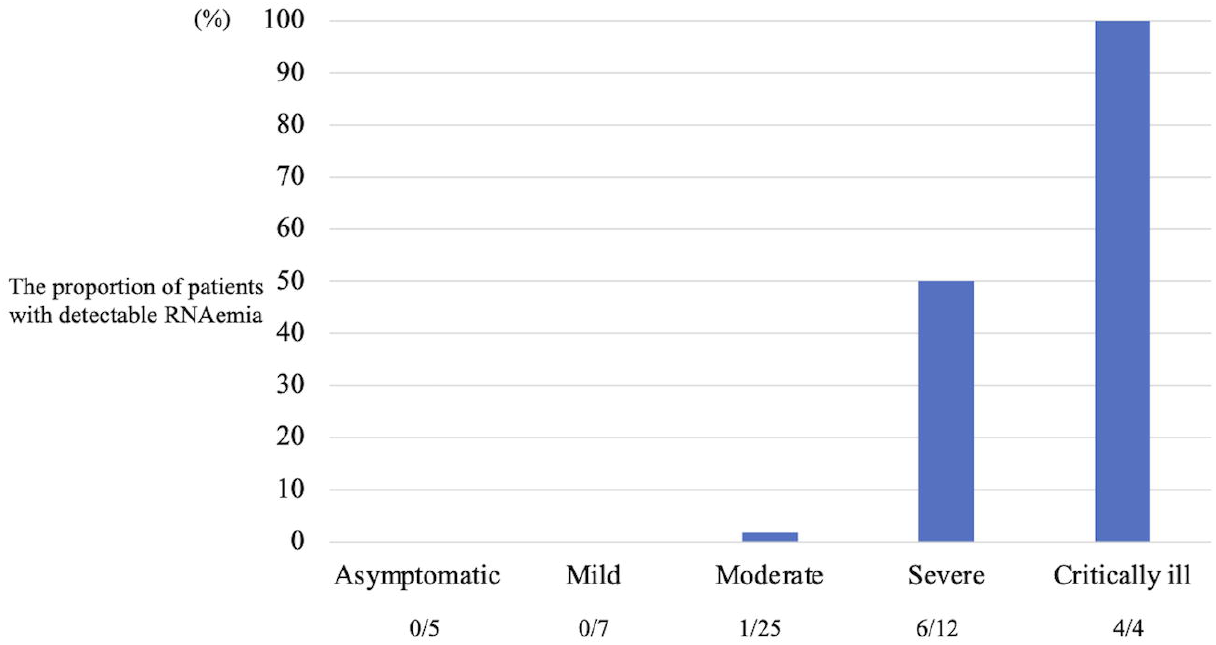
The proportion of the patients with detectable RNAemia stratified with severity.

For their clinical outcomes, patients with RNAemia required more frequent oxygen supplementation (90.0% vs. 13.3%; *P* < .0001) and ICU admission (81.8% vs. 6.7%; *P* < .0001) and required invasive mechanical ventilation (27.3% vs. 0.0%; *P* < .0001). In-hospital mortality was more frequent in patients with SARS-CoV-2 RNAemia than in those without SARS-CoV-2 RNAemia; however, this difference was not significant (18.2% vs. 2.2%; *P* =.095) possibly because of the small sample size (Table 2).

**Table 2.**
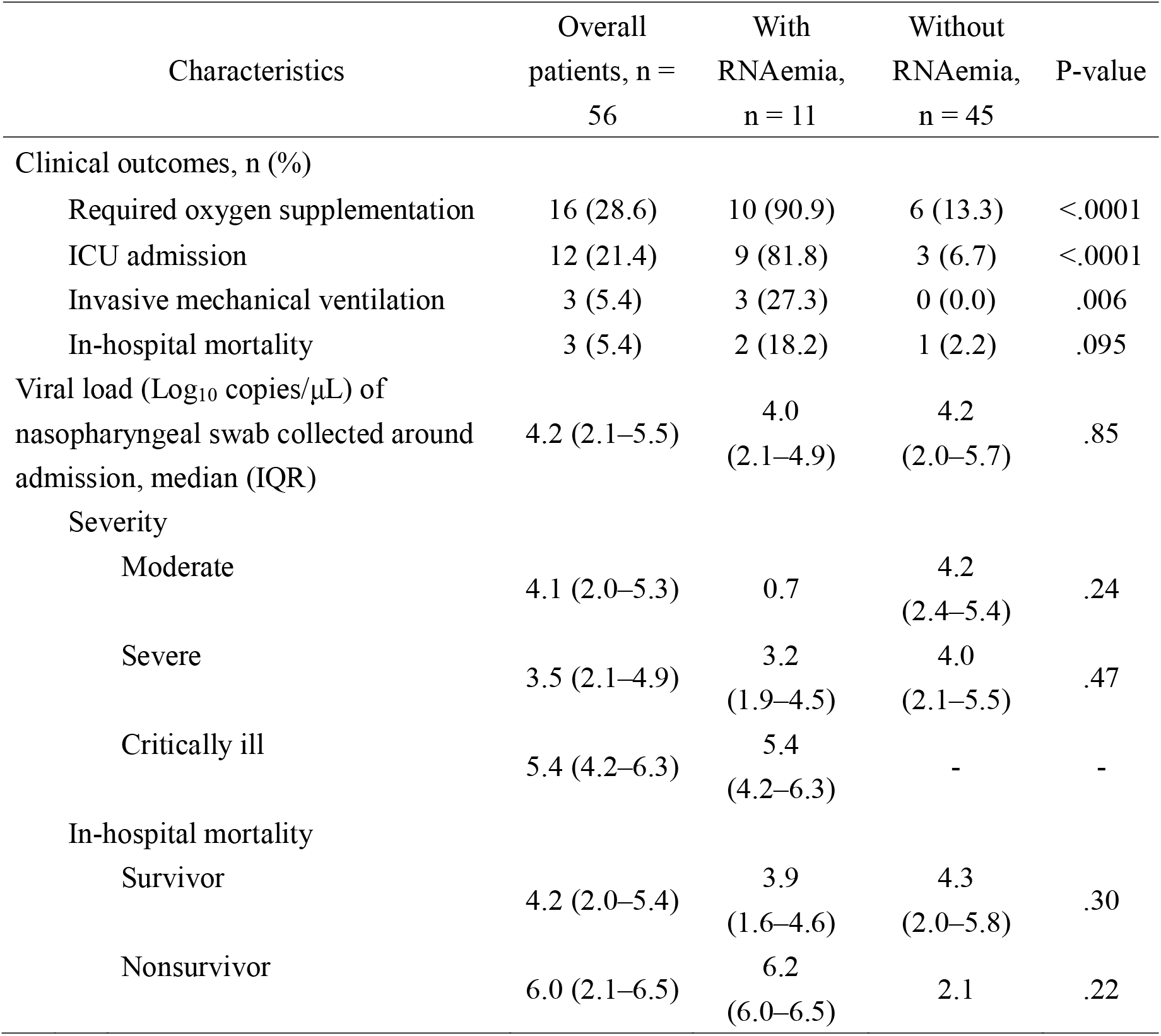
COVID-19 clinical severity outcomes and nasopharyngeal viral load association with detectable RNAemia in 56 patients.

| Characteristics | Overall<br>patients, n =<br>56 | With<br>RNAemia,<br>n = 11 | Without<br>RNAemia,<br>n = 45 | P-value |
| --- | --- | --- | --- | --- |
| Clinical outcomes, n (%) |  |  |  |  |
| Required oxygen supplementation | 16 (28.6) | 10 (90.9) | 6 (13.3) | <.0001 |
| ICU admission | 12 (21.4) | 9 (81.8) | 3 (6.7) | <.0001 |
| Invasive mechanical ventilation | 3 (5.4) | 3 (27.3) | 0 (0.0) | .006 |
| In-hospital mortality | 3 (5.4) | 2 (18.2) | 1 (2.2) | .095 |
| Viral load (Log<sub>10</sub> copies/μL) of<br>nasopharyngeal swab collected around<br>admission, median (IQR) | 4.2 (2.1–5.5) | 4.0<br>(2.1–4.9) | 4.2<br>(2.0–5.7) | .85 |
| Severity |  |  |  |  |
| Moderate | 4.1 (2.0–5.3) | 0.7 | 4.2<br>(2.4–5.4) | .24 |
| Severe | 3.5 (2.1–4.9) | 3.2<br>(1.9–4.5) | 4.0<br>(2.1–5.5) | .47 |
| Critically ill | 5.4 (4.2–6.3) | 5.4<br>(4.2–6.3) | - | - |
| In-hospital mortality |  |  |  |  |
| Survivor | 4.2 (2.0–5.4) | 3.9<br>(1.6–4.6) | 4.3<br>(2.0–5.8) | .30 |
| Nonsurvivor | 6.0 (2.1–6.5) | 6.2<br>(6.0–6.5) | 2.1 | .22 |

There was no significant difference in viral loads in the NP swabs of patients with RNAemia (4.0 Log_10_ copies/μL [IQR, 2.1–4.9]) and without RNAemia (4.2 Log_10_ copies/μL [2.0–5.7]; *P* = .85) (Table 2). The overall median number of days of symptoms at the time of NP testing was six days (IQR: three to nine) and was similar in moderately to critically ill patients with and without RNAemia.

In an in-depth study of RNAemia patients, although the days after symptom onset at the time of NP testing were not significantly different, the median viral loads of NP swabs were significantly higher in critically ill cases (5.4 Log_10_ copies/μL [IQR, 4.2–6.3]) than in moderate–severe cases (2.6 Log_10_ copies/μL [1.1–4.5]; *P* =.030) and in nonsurvivors (6.2 Log_10_ copies/μL [IQR, 6.0–6.5]) than in survivors (3.9 Log_10_ copies/μL [1.6–4.6]; *P* = .045).

## Discussion

This analytical cross-sectional study assessed 56 individuals for the detection of RNAemia in serum samples. These findings show that RNAemia was present in almost 20% of the samples tested. The results from other studies show discordant rates of SARS-CoV-2 detection in serum (ranging from 10.4% to 74.1%) (1, 7–11), whereas other authors did not find any patients or reported only 1% of RNAemia (1, 7, 8). However, a direct comparison of these results is challenging because of the use of different testing protocols, including differences in specimen extraction volumes (ranging from 80 to 200 μL), sample types examined (plasma, serum, and whole blood), viral genomic targets tested (N, S, E genes or ORF1ab region), variable timing of blood collection relative to symptom chronology, and different patient populations (9).

Regarding the clinical significance of the presence of SARS-CoV-2 RNAemia, our results agree with those reported by other authors, thus suggesting an association with ICU admission, invasive mechanical ventilation, and mortality (11, 13–15). On the other hand, the association between viral load in respiratory specimens and severity remains unclear and controversial in the literature. Multiple case series reported that the viral load of asymptomatic patients is as high as that of asymptomatic patients (3, 10). Meanwhile, recent studies found that NP viral load at admission independently correlates with the risk of intubation and in-hospital mortality (11). This discrepancy may be attributed to different patient populations, including severity levels.

In this study, there was no significant difference in viral load in the NP swabs of patients with and without RNAemia. These results confirm those from Berastegui-Cabrera et al. (1) and Prebensen et al. (12), who did not find an association between the viral load in NP samples and the presence of SARS-CoV-2 RNAemia or correlation with the viral load in blood. However, among the patients with RNAemia in the present study, the higher viral load of NP swabs was correlated with the severity and mortality. This result suggested that NP viral load may also be useful as an aid for the more accurate prediction of the severity and mortality of patients with RNAemia than RNAemia alone.

Although this study was not performed in a controlled setting, most serum samples with the presence of RNAemia and NP samples in mild to critically ill patients were collected before or on the same day as their admission, thus supporting a possible role for the early detection of at-risk individuals. Thus, in addition to assessing the viral load of NP swabs, which is the technique that is most frequently used for COVID-19 diagnosis, serum tests may be considered a complementary modality for the early identification of individuals who are likely to develop severe COVID-19.

This study has several limitations inherent to its small sample size, retrospective design, and potential for confounding the detection of SARS-CoV-2 RNA from the serum, NP viral load, and clinical conditions that cannot be excluded. In addition, we did not conduct multivariate and age-adjusted analyses owing to the strong multicollinearity between variables and the small sample size. Longitudinal analyses were also not performed because viral load dynamics could be modified by the received treatment, including combinations of antivirals and antibiotics during hospitalization.

## Conclusions

In summary, this study demonstrated a relatively high overall proportion of detectable SARS-CoV-2 RNAemia, which is an association between serum viral detection and older age, and severe clinical disease, including the need for oxygen supplementation and ICU admission, invasive mechanical ventilation, and likely in-hospital mortality. In addition, among the patients with RNAemia, the viral loads of NP swabs were significantly higher in severe and nonsurvivor cases than in non-severe and survivor cases. Further studies are required to investigate the prognostic potential of RNAemia and those combined with the viral load of NP swabs to facilitate early therapeutic and supportive interventions before the development of severe clinical manifestations of COVID-19.

## Data Availability

All relevant data are within the manuscript.

## Acknowledgment

We thank Ai Kakumoto for her support in COVID-19 testing.

